# A study of relationship between Social Determinant of Health and Imaging based Age Estimation using Head CT

**DOI:** 10.1101/2023.05.27.23290611

**Authors:** Amara Tariq, Judy Gichoya, Bhavik N. Patel, Imon Banerjee

## Abstract

**Background:** The biological age of a person represents their cellular level health in terms of biomarkers like inflammation, oxidative stress, telomere length, epigenetic modifications, and DNA damage. Biological age may be affected by extrinsic factors like environmental toxins and poor diet indicating socioeconomic disadvantage. While biological age can provide a much more accurate risk estimate for age-related comorbidities and general decline in functioning than chronological age, it requires well-established laboratory tests for estimation.

**Methodology:** As an alternative to laboratory testing for biological age estimation, Incidental medical imaging data may demonstrate biomarkers related to aging like brian tissue atrophy. In this study, we designed a deep learning based image processing model for estimation of biological age from computed tomography scans of the head. We then analyzed the relation between gap in biological and chronological age and socioeconomic status or social determinants of health estimated by social deprivation index (SDI).

**Results:** Our CNN based image processing regression model for biological age estimation achieves mean absolute error of approximately 9 years between estimated biological and chronological age with -0.11 correlation coefficient with SDI. With the fusion of imaging and SDI in the process of age estimation, mean absolute error is reduced by 11%.

**Conclusion:** The results of our experiments clearly establish a correlation between social determinants of health and the gap between biological and chronological ages.

## Introduction

While chronological age is based on time elapsed from the time of the birth of a person, cellular or biological age (BA) is a measure of gradual decline in cell division and proper functioning. BA may be affected by intrinsic factors like genetic changes and cellular metabolism as well as extrinsic factors like exposure to environmental toxins, lifestyle, and diet[1-3]. While chronological age is often used as a crude estimate of BA to assess risk of mortality and morbidity, BA can provide a more accurate risk estimate. Even with lack of a universally agreed upon definition and quantification of BA, laboratory test based BA estimation measures have been well-established[4-6].

Biological age related markers may be incidentally collected in medical data such as physical activity from wearable devices[7,8] and medical imaging data[9-16]. Age-related changes in bone density [17, 18], body composition [19-21], cardiovascular structures [22, 23], lungs [24, 25], and ocular structures [26, 27] have been demonstrated on imaging. Deep learning based image processing models have recently been designed to quantify such biomarkers for biological age estimation using magnetic resonance imaging of the brain [28, 29], retinal photographs [30, 31], and chest X-rays [32]. “*Brain age*” has been established as indicative of cognitive decline and mortality[28, 29]. Degree of atrophy in brain tissue captured in radiological images such as brain MR and head CT, can indicate biological age of a person which may be different from chronological age.

Biological age estimation studies have often connected risk of mortality and morbidity with larger gaps between chronological and biological age[30, 31]. While this gap may be caused to some extent by intrinsic factors, social determinants of health (SDoH) can influence biological age, accelerating cellular aging and overall health outcomes [33]. Electronic health records (EHR) collected for patients in hospitals may provide clues to social determinants of health but cannot guarantee quantification of its influence. Objective of the current study is to understand the relation between biological age calculated using imaging features calculated from head CT studies and social determinants of health. Our hypothesis is that higher biological age than chronological age primarily represents poor quality of life including unhealthy diet, stress, exposure to environmental toxins, sedentary lifestyle, etc.

Social deprivation index (SDI) is a composite measure of seven demographic characteristics collected in the American Community Survey (ACS) to quantify social determinants based on geolocation: percent living in poverty, percent with less than 12 years of education, percent single-parent households, the percentage living in rented housing units, the percentage living in the overcrowded housing unit, percent of households without a car, and percentage nonemployed adults under 65 years of age. SDI can address challenges in quantifying social determinants of health to best guide clinical and community health interventions.. The SDI measure was calculated at the four geographic areas: county, census tract, aggregated Zip Code Tabulation Area (ZCTA), and Primary Care Service Area (PCSA, v 3.1). While raw values are directly computed by the formula using the seven measures described above, SDI score is a value normalized between 0-100 with higher value indicating higher extent of social disadvantage. We use SDI score as a surrogate for social determinant of health and study the relation between gaps in biological age estimated by our deep learning based imaging model and chronological age of the patient at the time of the imaging exam. In order to prove our hypothesis, we adopted two parallel experiments - (i) (post-processing) establish the correlation between model computed biological age and SDI; (ii) (in-processing) include SDI during the learning of the model which may boost the model performance for biological age by providing surrogate for social determinant of the patient..

## Cohort selection

With the approval of Mayo Clinic Institutional Review Board (IRB), we included 3875 non-contrast head CT exams (sagittal views) with no acute finding performed between 2015-2022 belonging to 2433 patients. Patients were randomly split into training, test and validation sets. Table 1 shows demographic features of each split of the cohort. Patients’ chronological ages were computed through birth dates and head CT exam dates recorded in electronic health records (EHR). 9-digit zip codes were also extracted from EHR from the recorded addresses of patients.

**Table 1:**
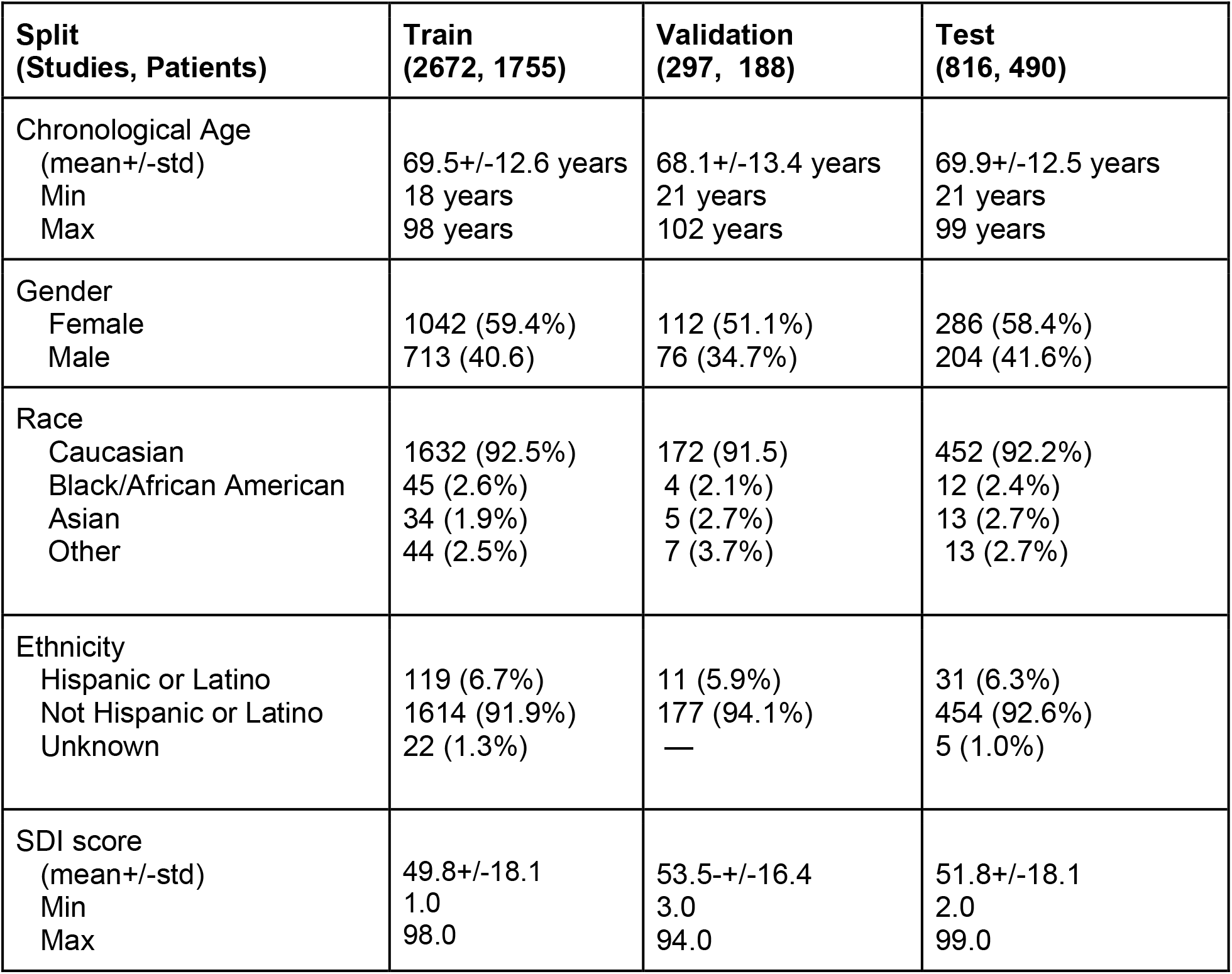
Cohort statistics

| **Split**<br>**(Studies, Patients)** | **Train**<br>**(2672, 1755)** | **Validation**<br>**(297, 188)** | **Test**<br>**(816, 490)** |
| --- | --- | --- | --- |
| Chronological Age<br>(mean+/-std)<br>Min<br>Max | 69.5+/-12.6 years<br>18 years<br>98 years | 68.1+/-13.4 years<br>21 years<br>102 years | 69.9+/-12.5 years<br>21 years<br>99 years |
| Gender<br>Female<br>Male | 1042 (59.4%)<br>713 (40.6) | 112 (51.1%)<br>76 (34.7%) | 286 (58.4%)<br>204 (41.6%) |
| Race<br>Caucasian<br>Black/African American<br>Asian<br>Other | 1632 (92.5%)<br>45 (2.6%)<br>34 (1.9%)<br>44 (2.5%) | 172 (91.5)<br>4 (2.1%)<br>5 (2.7%)<br>7 (3.7%) | 452 (92.2%)<br>12 (2.4%)<br>13 (2.7%)<br>13 (2.7%) |
| Ethnicity<br>Hispanic or Latino<br>Not Hispanic or Latino<br>Unknown | 119 (6.7%)<br>1614 (91.9%)<br>22 (1.3%) | 11 (5.9%)<br>177 (94.1%)<br>— | 31 (6.3%)<br>454 (92.6%)<br>5 (1.0%) |
| SDI score<br>(mean+/-std)<br>Min<br>Max | 49.8+/-18.1<br>1.0<br>98.0 | 53.5+/-16.4<br>3.0<br>94.0 | 51.8+/-18.1<br>2.0<br>99.0 |

## Methodology

### Imaging model for age prediction

Within the scope of this study, we used non-contrast CT studies with ‘no acute finding’ and extracted the middle slice from the sagittal direction from the axial thins (<1mm slice thickness). The images are processed with application of soft-tissue window (50, 100). To predict the numeric age, these slices were fed through a DenseNet-121 based architecture with classification layer substituted with regression layer. The model was trained to minimize mean squared error (MSE) computed by comparing model output with normalized value of chronological age computed through birth dates recorded in electronic health records (EHR).

For analysis of discrepancy in predicted and chronological age, the normalized predicted output of the model was mapped back to actual age range (between 20 and 100 years). In addition, 9-digit zip code recorded in EHR for each patient was mapped to county identifiers form Federal Information Processing System (FIPS) through a zipcode-to-FIPS table collected from online resources made available by Center for Disease Control (CDC)^1^. Database of social deprivation index for each county was collected from Robert Graham Center^2^. Thus, SDI for the residential area of each patient in our database was collected. We estimated mean social deprivation scores for samples with larger than X years difference between estimated and chronological age, and varied X between 0 to +/-20 with negative values indicating that the age estimate was higher than chronological age and vice versa. We observed that social deprivation score and discrepancy between estimated and chronological age values are proportional to each other.

In line with this observation, we decided to infuse the age prediction model’s proceeding with social deprivation index to evaluate if the model’s predictions can be made unbiased to the socio-economic conditions of a patient’s residential area. Figure 1 shows the model architecture for our SDI-infused predictor inspired by DenseNet-121 architecture. Output of intermediate layers of the model are manipulated by SDI before being passed on the 4th denseblock and final classification layer. Our experiments indicate that the discrepancy in predicted and chronological age is no longer proportional to SDI score after this processing, in addition to achieving prediction performance improvement in terms of lower MAE.

**Figure 1:**
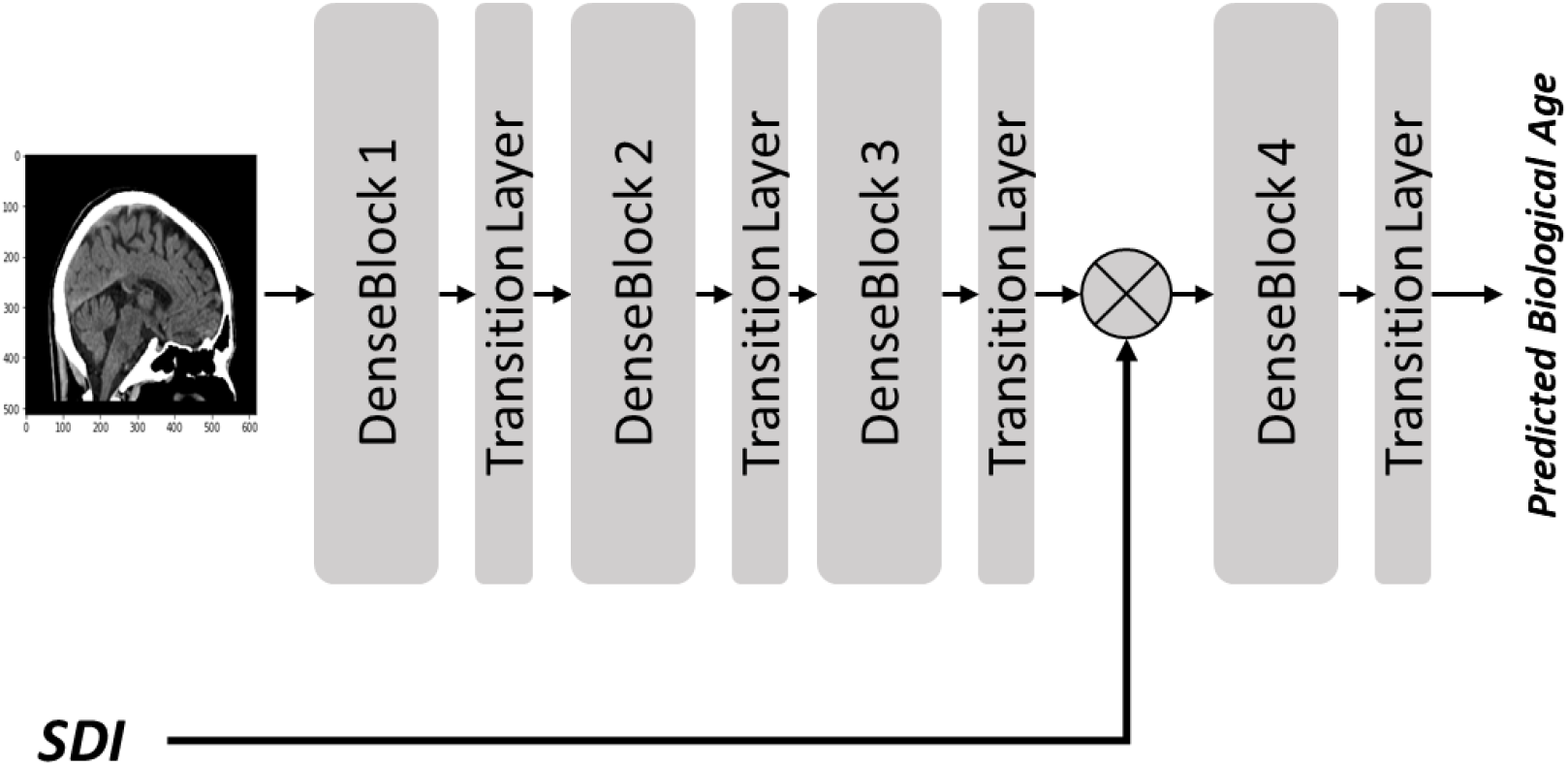
Fusion of SDI and brain CT image for age prediction

## Results

With an optimal learning rate of 1e-5 and 200 epochs of training, the model saturated to mean absolute error of 0.009 for validation data. When predicted values were mapped back to the chronological age range, we observed an average discrepancy of 8.8+/-6.8 years in the test set Figure 2 shows a scatter plot for predicted and chronological ages. This result highlights the limitation of age estimation from head CT imaging data. However, we argue that this limitation stems from biological and chronological age. While the model can only predict biological age, it is being evaluated against chronological age. We also argue that socio-economic factors affect the difference in biological and chronological age values. Taking the social deprivation score of the residential area of a patient as a surrogate for their socio-economic status and estimated age as a surrogate for biological age, we analyzed differences in biological and chronological age against social deprivation score. Figure 3 shows that patients with higher biological age than the chronological age come from areas with higher mean social deprivation score indicating poor quality of life compared to patients with biological age equal to or less than the chronological age.

**Figure 2:**
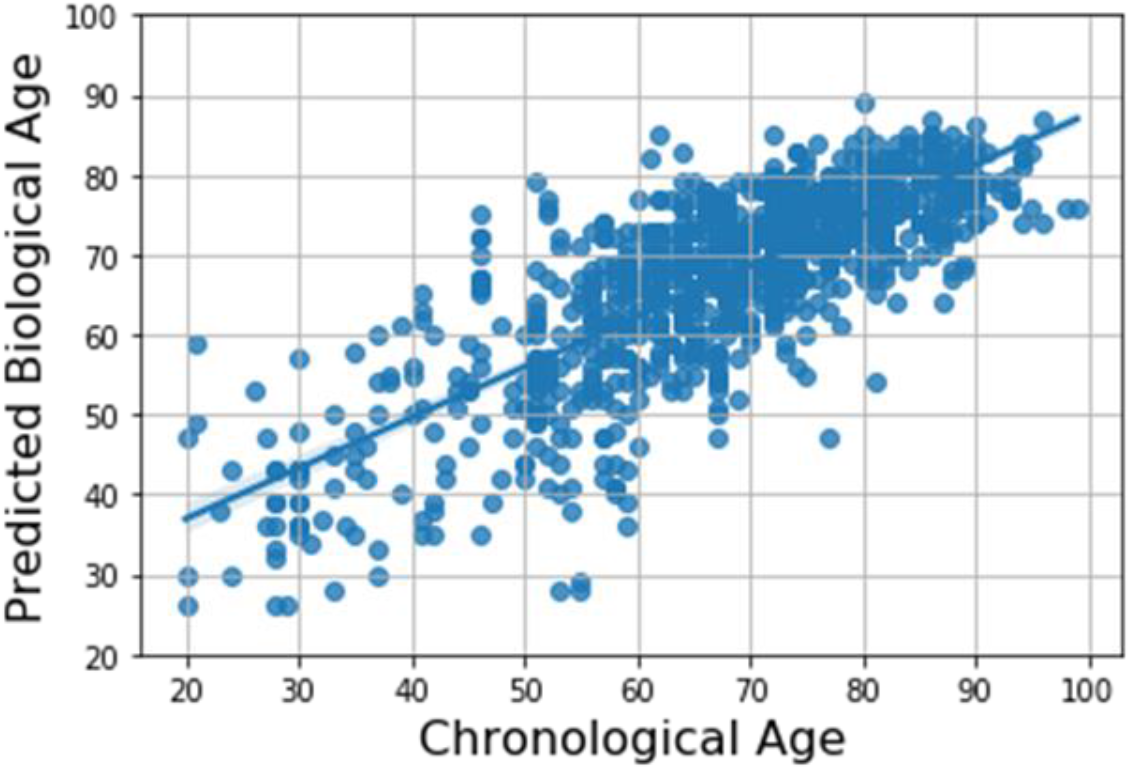
Scatter plot for predicted biological and chronological age

**Figure 3.**
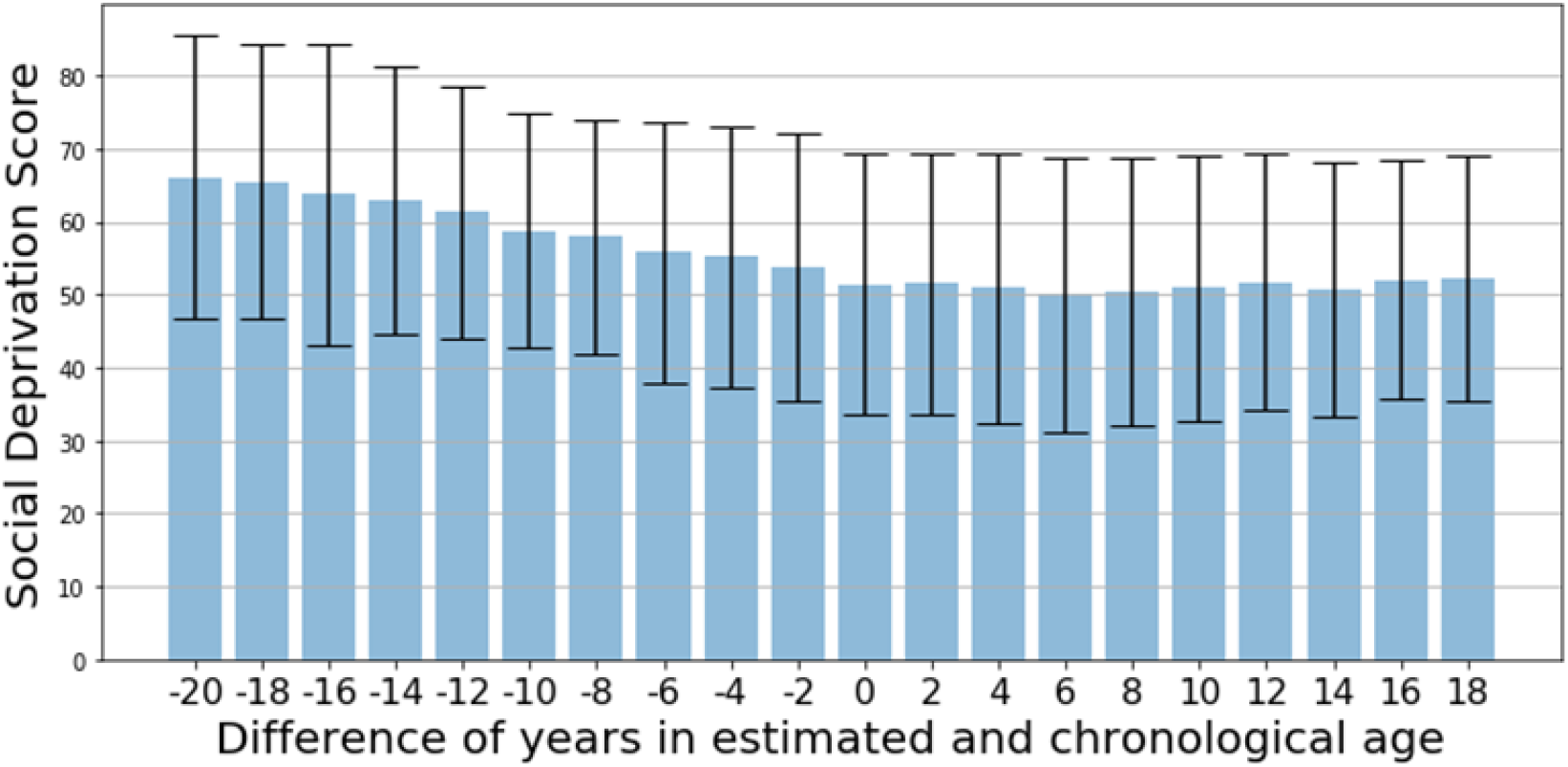
Difference in predicted biological and chronological age is proportional to SDI score. Higher SDI indicates poorer socio-economic conditions of the area. For patients with biological age higher than chronological age, mean SDI scores are higher than patients with biological age lower than chronological age.

When social deprivation index is infused into the model, this correlation trend between predicted age (surrogate of biological age) and chronological age vanishes as indicated by Figure 4 and Pearson correlation coefficient comparison reported in Table 2. Marginal performance improvement is also observed in terms of lower MAE of 7.9+/-6.3 year.

**Figure 4.**
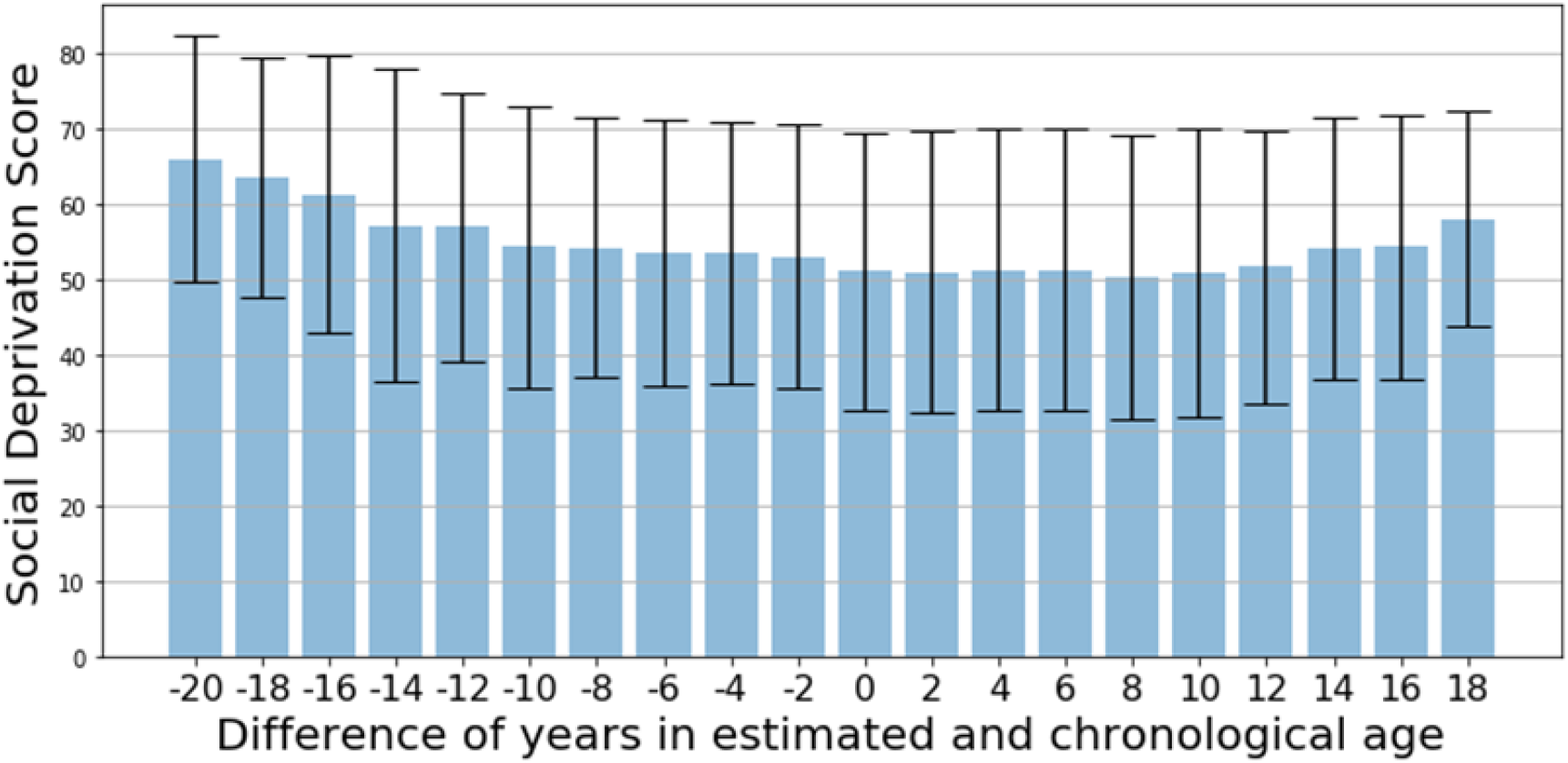
For SDI-infused biological age predictions, difference in predicted biological and chronological age is no longer proportional to SDI score.

**Table 2:** Age prediction performance for models with and without sdi

| Biological Age Estimator | Pearson correlation coefficient between age gap and SDI | Mean Absolute Error mean+/-std (in years) |
| --- | --- | --- |
| Head CT | -0.11 | 8.8+/-6.8 |
| Head CT and SDI | -0.07 | 7.9+/-6.3 |

We analyzed the model’s output by using gradients of the target values passing from convolutional layers to produce a localization map highlighting important regions in the image for target value prediction (Figure 5). It is evident that the model is focusing on texture in the brain region of the image while making predictions.

**Figure 5:**
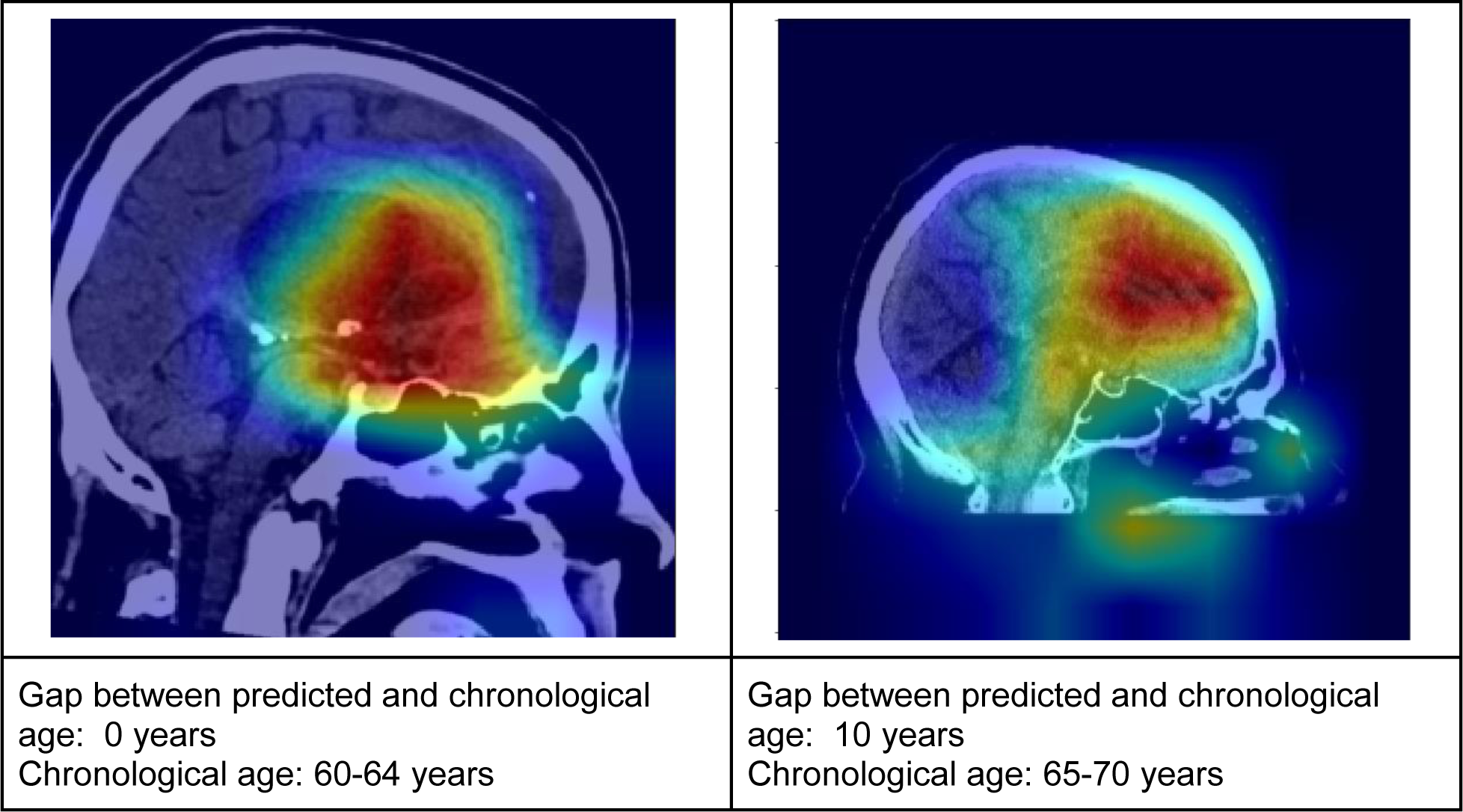
Heatmap indicates model-assigned importance to image regions for age estimation

In comparative experiments, we noticed that when SDI-infused prediction is closer to chronological age, the infusion of SDI seems to have been able to focus the model’s attention brain area earlier in the processing layers (3rd denseblock compared to the 4th denseblock) as compared to the model without SDI. Figure 6 shows the processing of the same sample with and without SDI where prediction of model with SDI is closer to chronological age compared to model without SDI.

**Figure 6:**
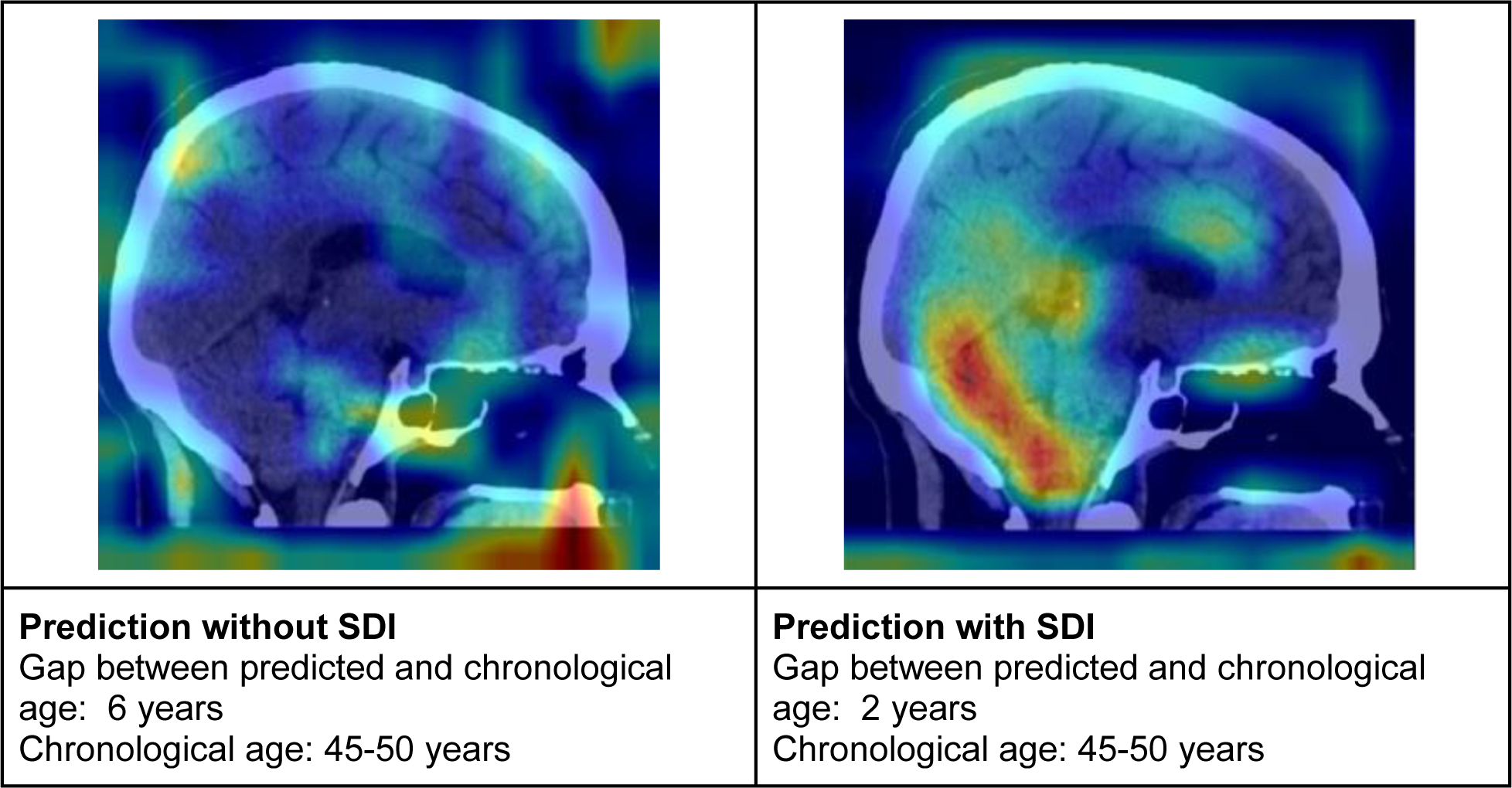
Heatmaps for model-assigned importance to image regions for age estimation with (right) and without (left) SDI.

## Discussion

Cellular or biological age of a person is indicative of health of a person at cellular level indicated by biomarkers like inflammation, oxidative stress, telomere length, epigenetic modifications, and DNA damage. Biological age can identify patients at higher risk of age-related diseases or functional decline. However, the current assessment of biological age is limited by the absence of a universally accepted approach for assessment and the invasive nature of the testing methods employed[34, 35]. An alternative approach is to quantify established biomarkers demonstrated in medical imaging data such as brian CT and MR and retinal images[28-32]. Often such quantification of biological age is accompanied by an analysis of correlation between biological age and mortality and morbidity

While biological age is demonstrated by intrinsic factors like cellular health, extrinsic factors like exposure to environmental toxins, quality of diet, and lifestyle have a sizable impact on cellular health. Such extrinsic factors are often expressed as social determinants of health. For design of community-level healthcare intervention as well as personalized assessment of age-related comorbidities, it is important to understand the connection between biological age and social determinants of health. This is the main goal of this study.

We first design a predictive model that can estimate the biological age of the patient through deep learning based processing of sagittal view non-contrast enhanced head CT. The limitation of this model development was the fact that we only had access to the patient’s chronological age values during the training process. Hence, the model, while only capable of estimating biological age, was penalized for discrepancy in predicted and chronological age. Due to this unique scenario, we were more interested in understanding the error of the model’s predictions, i.e., the difference between predicted biological and groundtruth chronological age rather than traditional quality measures for regression models like lean absolute error (MAE). We hypothesized that the gap between estimated biological and chronological age is connected with social determinants of health. We used the social deprivation index as a surrogate measure for gauging quality of life or relative socioeconomic disadvantage. Hypothetically, patients with larger socioeconomic disadvantage, as indicated by higher SDI, would have a larger negative gap between estimated biological and chronological ages (with estimated biological age higher than chronological age). Our hypothesis was confirmed through experimental results.

We also attempted to fuse social determinants of health in age estimation through design of densenet inspired architecture where processed representations of imaging data are manipulated with corresponding SDI values before prediction of final output. As expected, the ‘error’ in new model’s outputs, i.e., the difference in biological and chronological age were no longer directly affected by SDI values as the model had incorporated SDI already in the process of output prediction which loss function tried to push closer to chronological age as explained in the previous section. However, little improvement was recorded in overall regression quality in terms of mean absolute error. This may be caused by the limitation of the study that head CT were collected from patients presenting at a large academic healthcare institute where a much higher portion of population lies in 50 to 70 years age range rendering estimation of age outside of this range considerably harder.

## Data Availability

All data produced in the present study are available upon reasonable request to the authors

## Footnotes

1 https://wonder.cdc.gov/wonder/sci_data/codes/fips/type_txt/cntyxref.asp

2 https://www.graham-center.org/maps-data-tools/social-deprivation-index.html

